# Low cost, injection molded, nasopharyngeal swabs for addressing global diagnostic supply shortages

**DOI:** 10.1101/2021.07.19.21260235

**Authors:** Manuel Ramses Martinez, Rachelle Mendoza, David Perry, Elmer Gabutan, William Fyke, Eve Faber, Subodh J Saggi, Jenny Tam, Isabel Chico-Calero, Heather Centner, David M. Engelthaler, Laura H. Goetz, Timothy K. McDaniel, Noriyuki Murakami, Jenny Libien, Donald E. Ingber, Richard Novak

## Abstract

The SARS-CoV-2 pandemic revealed the fragility of global supply chains, resulting in shortages of essential clinical supplies for the detection and treatment of COVID-19, including nasopharyngeal swabs (NP) commonly used to collect clinical samples for respiratory disease diagnostics. Groups around the world led a response to find alternatives to these shortages. Here, we describe the development of a single-component nasopharyngeal swab (GrooveSwab) composed of medical grade polypropylene that can be mass manufactured using conventional injection molding. The design inspired by cat tongue papillae consists of a non-absorbent stacked ring structure that allows for efficient and comfortable collection of viscous mucus samples and faster release into collection medium compared to traditional absorbent swabs. GroveSwabs were evaluated in 2 clinical studies in the US and Peru and demonstrated 90% or better concordance with commercial swabs. The simplified NP swab offers a reliable, lower cost, and more manufacturable alternative to absorbent swabs to avoid future shortages.

## INTRODUCTION

Since December 2019, severe acute respiratory syndrome coronavirus 2 (SARS-CoV2) and the associated COVID-19 pandemic has spread across the globe, infecting millions of diagnosed individuals to-date^1^ and disrupting global supply chains. Nasopharyngeal (NP) swabs have been considered the gold-standard means of collecting specimens for the diagnosis of respiratory virus infections^2–5^. These specialized swabs consist of a handle, a flexible neck, and a head coated in short bristles (flock) or absorbent foam. Due to their specific geometry and other design criteria, NP swabs have been particularly difficult to obtain, and numerous efforts have focused on alternative sampling sites^2,4,6,7^. The global shortage of medical supplies spurred multiple efforts to develop alternative NP swab solutions, including 3D printed swabs^8–11^, which do not require complex, multi-step manufacturing. The flexibility of 3D printing facilitated a rapid response to sudden swab shortages; however, the relatively high cost of these swabs and lack of global availability of industrial scale 3D printing resources has limited the spread of the technology.

Injection molding is the standard manufacturing modality for many medical products, and the infrastructure for injection molding exists globally and it offers the lowest cost per part at large production scales. Here we leverage established medical device manufacturing using single-shot injection molding and a single-material design to facilitate the development of low-cost nasopharyngeal swabs with the functionality of a traditional flocked or foam-tipped NP swab, but using a simpler manufacturing process and allowing greater availability and versatility of use in the clinic.

## MATERIALS AND METHODS

### Swab design and manufacturing

The GrooveSwab NP swabs were designed in SolidWorks (Dassault Systèmes) incorporating input criteria from clinicians, including those previously published^8^. Supplemental **Figure S1** describes the design geometry. Swabs were injection molded from medical grade P5M4R-034 polypropylene (Flint Hills Resources) using rapid prototyping tooling from Protolabs. Swabs were sterilized by autoclave (121°C, 20 min) in autoclave-compatible pouches. For clinical trials, swabs were manufactured under ISO13485 conditions by Innovative Product Brands, Inc. (Highland, CA, USA) and packaged singly in peel pouches and sterilized using ethylene oxide. Innovative Product Brands, Inc. also manufactured swabs with polyester fiber flocked heads using electrostatic fiber alignment and adhesion.

### Fluid dynamics simulation

The finite element simulation of fluid dynamics using the swabs was performed using the fluent module of the Ansys software package (Ansys, Canonsburg, PA). Higher density meshing was used in the regions surrounding the swab ring structures to improve simulation of the wall shear and velocity magnitude gradients. In addition, dynamic mesh functions were used to adapt the mesh according to the displacement of the swab. Wall shear and velocity magnitudes were calculated during the application of a sinusoidal displacement of the swab inside a 6.8 mm inner diameter vial (e.g., ThermoFisher Matrix Tube), with an amplitude of 15 mm/s and a period of 4 seconds. Water was used as the working fluid and was solved using the standard k-epsilon turbulence model. The simulation was performed in a transient state of 4 s to observe the behavior of the fluid as a function of the swab displacement in the vial.

### Mock sample collection and release analysis in vitro

A mock mucus solution was prepared using 3% w/v porcine gastric mucin (Sigma-Aldrich) in phosphate buffered saline (PBS)^12^, and fluorescent inulin-FITC was added to the solution at 0.1 mg/mL from a 5% w/v stock solution and mixed thoroughly. Swab heads were inserted for 10 s and rotated to mimic CDC collection guidelines and removed along the tube wall. Swabs were placed into vials containing 3 mL PBS and removed immediately or after various timepoints. 100 μL of well-mixed eluate was pipetted into 96 well plates, and fluorescence was measured using a BioTek NEO2 plate reader configured for FITC fluorescence.

### Clinical sample collection and analysis

The TGen North study comparing the prototype GrooveSwab against a Copan FLOQSwab (Copan Diagnostics, Murietta, CA, USA) involved the retrospective analysis of anonymous human data that had originally been generated in validation testing as part of a clinical laboratory quality assurance program. The original samples were collected from healthy volunteers who were participating in a workplace surveillance testing program. The retrospective analysis of the data was determined to be non-human subjects research by the TGen Office of Research Compliance & Quality Management.

To enable testing of potential positive cases in a healthcare environment, patients at SUNY Downstate (Brooklyn, NY, USA) were recruited for comparing the prototype GrooveSwab against a control swab, the Puritan PurFlock Ultra (Puritan, Guilford, ME, USA) under IRB protocol number 1611734-2. Consented patients consisted of a mix of symptomatic and asymptomatic patients in outpatient, Emergency Department, and inpatient settings. Patients known to be COVID-19 positive, including patients recovering from COVID-19, were included in the study. Medical personnel performing the swabbing and patients were asked to evaluate their experience using a subjective preference survey that explored ease of use and comfort related to the sample collection process. Collection followed CDC guidelines^13^ for NP swab sample collection. Briefly, the swab head was inserted through one nostril parallel to contact the nasopharynx. Swabs were rotated, brushed against the nasopharynx several times, left in place for several seconds and then removed. Following removal from patient, the swab head was placed in a vial containing 3 mL of viral transport medium (BD universal viral transport system, Franklin Lakes, NJ, USA) or 0.9% saline and the handle was broken off by 1) bending it 90 degrees at the midpoint notch, followed by 2) twisting the handle for a complete break. A control swab was used in the opposite nostril, with swabbing performed in a similar manner.

At TGen North, immediately following collection, specimens were placed into a sterile tube containing 3mL phosphate buffered saline. Each tube was briefly vortexed and stored at 4ºC for no longer than 72 hours prior to processing. RNA was extracted from samples using the Quick-RNA Viral Kit (Zymo Research, Irvine, CA), following the manufacturer’s protocol. RT-PCR was performed using Reliance One-Step Multiplex RT-qPCR Supermix (Bio-Rad, Hercules, CA) in conjunction with TGen-developed primers for the SARS-CoV-2 N and S genes, and the CDC’s human RNase P primers as an internal positive control for RNA extraction on the CFX Connect instrument (Bio-Rad, Hercules, CA). All samples yielded a Ct value of less than 35 in the CDC RNase-P assay, indicating a valid test, and were negative for both SARS-CoV-2 viral assays (Ct> 40).

At SUNY Downstate, tubes with collected samples were inverted 5-6 times prior to obtaining 100-μl aliquot for viral RNA extraction. RNA extraction was conducted using the MagMAX Viral/Pathogen Nucleic acid isolation kit automated on the KingFisher Flex Purification system (Thermo Fisher Scientific, Waltham, MA) following manufacturer’s protocol. The RT-PCR, including cDNA synthesis and PCR amplification of the target sequences, was performed in either triplicate or duplicate PCR reactions using the QuantStudio 6 Flex Real-Time PCR system (Applied Biosystems, Foster City, CA). The assay utilized the primer and probe sets included in the TaqPath COVID-19 Combo Kit (FDA EUA on March 13, 2020) (Thermo Fisher Scientific, Waltham, MA). These primer and probe sets were designed to amplify and detect three regions of the SARS-CoV-2 single stranded RNA genome: the ORF1ab, N and S genes. In the same assay, MS2 phage RNA was used as internal positive control for RNA extraction and RT-PCR. The detection of 2 or more viral gene targets with cycle threshold (Ct) values of less than or equal to 37 indicated a positive result, whereas detection of only 1 of the three viral gene targets was deemed inconclusive and would be repeated once using the extracted RNA. MS2 amplification was required for the result to be considered valid. Some of the standard swabs were tested for COVID-19 using Cepheid GENExpert, Roche Cobas, or BioFire COVID-19 tests according to manufacturer instructions.

### Statistical analysis

Unless otherwise noted, paired t-tests were performed in Graphpad Prism with a p < 0.05 significance threshold. Concordance was calculated using Cohen’s kappa and by percent positive agreement.

## RESULTS

Our initial design was inspired by the ability of cat tongue papillae to capture fluids with high efficiency based on their shape and microfluidic behavior.^14^ Following prototyping using 3D printed nylon, the swabs that we termed GrooveSwabs were injection molded from medical grade FHR P5M4R polypropylene (**Fig. 1A**). The material was selected to be compatible with autoclaving (121°C, 20 min), ethylene oxide, gamma radiation, and e-beam sterilization. The stacked rings of the head (**Fig. 1B**) enable mucus collection from the nasopharynx without the need for a fibrous or flocked coating. A flat region was included in the head for robust demolding using an ejection pin without causing excess flashing or vestiges in areas of the head that would be exposed to contact with the nasal cavity. The flexible tapered neck and rounded tip of the swab head reduce likelihood of damage to the nasal cavity and nasopharynx, and deflection force was designed to be comparable to a Copan swab per clinicians’ guidance. Additional safety testing consisted of validating that while holding the swab handle, the tip of the swab could deflect >90 degrees off axis and be further rotated 90 degrees without fracturing or breaking to safely exceed typical use case conditions. The polypropylene material’s toughness reduces the likelihood of injury to patients while still enabling break-off of the handle for depositing the swab head into a collection vial.

**Figure 1.**
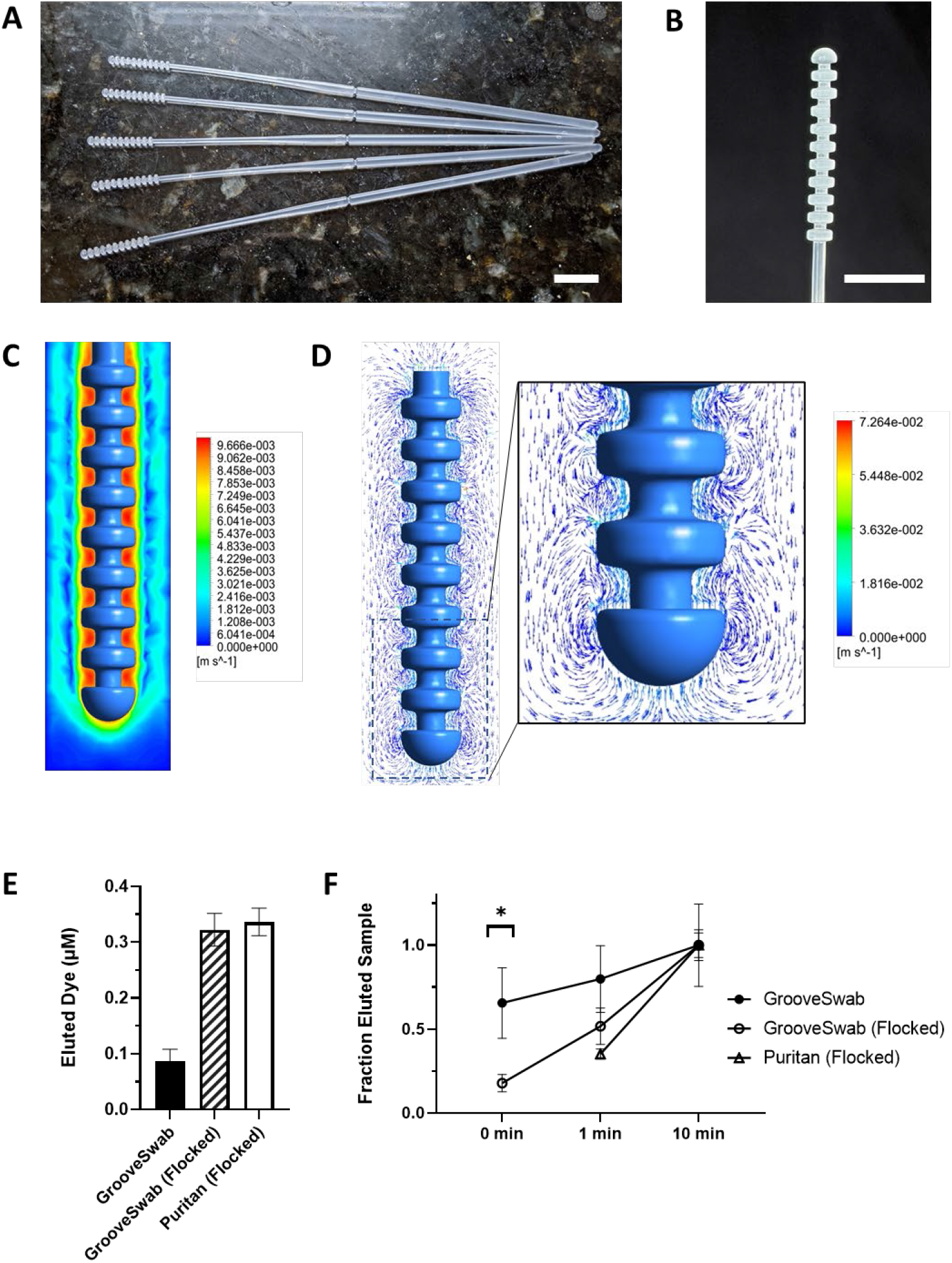
Photographs of A) injection-molded swabs B) swab head showing the stacked ring structure. Fluid dynamics simulation showing C) fluid velocity and D) velocity vectors highlighting vortex flow patterns as the swab is removed from a 6.5 mm inner diameter tube. Comparison of eluted mock respiratory mucus samples following 10 min elution in buffer (E) and over time (F).

Finite element simulation of fluid flow around a swab head during insertion and removal from a collection liquid indicated higher fluid velocity and shear rate around and in between the ring structures (**Fig. 1C,D**). The ability of swabs to collect mucus samples was first evaluated using a mock mucus fluid containing fluorescent dye. The GrooveSwab reproducibly collected approximately 1/3 as much material as a commercial flocked Puritan swab after 10 min incubation in buffer to release the sample; however, we obtained results comparable to the Puritan swab when we added flocking to the GrooveSwab design (**Fig. 1E**). More importantly, when we evaluated the smooth GrooveSwab head design’s ability to release collected mock samples into a collection buffer, we found that it released samples significantly faster in a single immersion step (0 min) compared to either flocked swab (**Fig. 1F**). Thus, given its simplicity and comparable performance, we used the non-flocked GrooveSwab in all subsequent studies.

The prototype non-flocked GrooveSwabs were tested in a “two nostril” test to assess sample collection efficiency. The prototype swabs were used together with a Copan FLOQSwab to test each healthy volunteer at the same time, with one swab inserted into one nostril and the second swab inserted in the other nostril immediately following the first. The test was administered by the same physician, and swabs were placed into sterile saline solution until RT-PCR testing later the same day. The Ct value for the RNAse P gene was used as a metric to assess the amount of cellular material collected by each swab. All swabs gave the expected negative test results: negative for both viral genes, positive for human RNAse P gene, used as a positive control to verify the collection of human cells by the swab. The prototype GrooveSwabs had a mean Ct value that was similar by slightly (0.8 cycles) higher on average than the Copan swabs (**Fig. 2A**).

**Figure 2.**
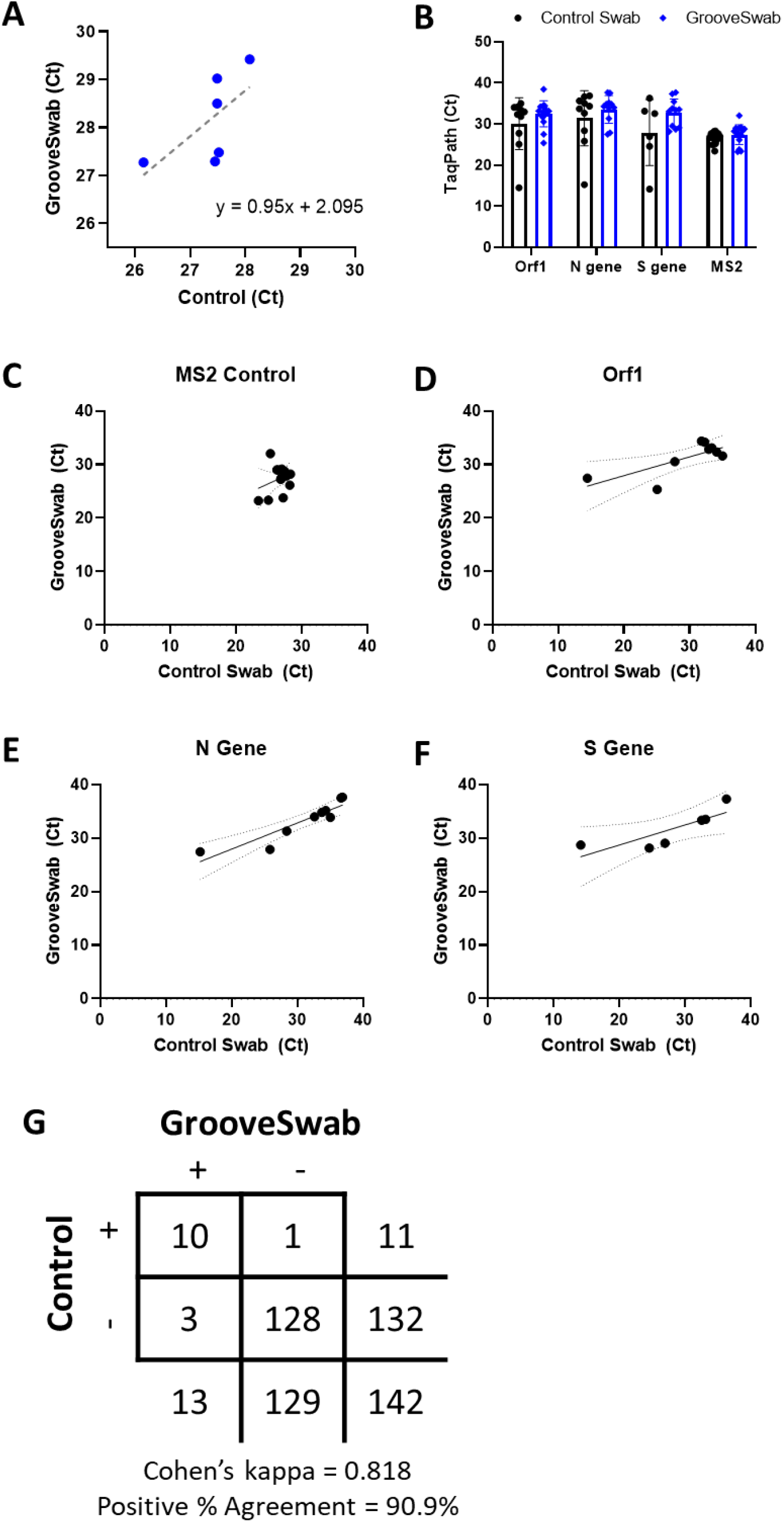
A) Comparison of RNAseP RNA between Copan FLOQswab and GrooveSwab from healthy volunteers. Ct values for three SARS-CoV2 target genes from positive patients comparing commercial flocked swabs against GrooveSwabs demonstrate agreement between swab groups (B) and direct correlation between swabs for each subject (C, D, E). G) The outcome of combined inpatient and outpatient study results showing 90% positive percent agreement.

To evaluate the performance of the prototype swabs in a clinical environment with patients presenting with COVID-19 symptoms and asymptomatic patients, we conducted concordance testing at SUNY Downstate Health Sciences University with patients seen in outpatient, Emergency Department, and inpatient settings. Subjects were swabbed by both GrooveSwab and Puritan swabs. Comparison of Ct values indicated a high degree of correlation across three viral genes (**Fig.2B-E**). A total of 146 patients participated in the study. Four patients’ results were determined to be inconclusive and were removed from further analysis. Of the 142 patients’ results that passed quality control, 10 tested positive by both swabs, and 1 by control swab only (**Fig. 2G**), resulting in Ct values not being significantly different and with substantial agreement (Cohen’s kappa = 0.818) with respect to the Puritan control swab. Additionally, 3 patients tested positive using the GrooveSwab but negative using the control swab. All 3 patients were confirmed COVID-19 hospitalized patients.

A subset (n = 58) of clinicians and patients were surveyed for usability and comfort, respectively. Clinicians reported roughly equivalent preference between the two swab types, with a slight preference for GrooveSwabs due to ease of gripping and of inserting and traversing nasopharynx (**Fig. 3A**). Subjective patient survey results indicated a strong preference for the GrooveSwabs (**Fig. 3B**) based on comfort during various phases of the sampling procedure.

**Figure 3.**
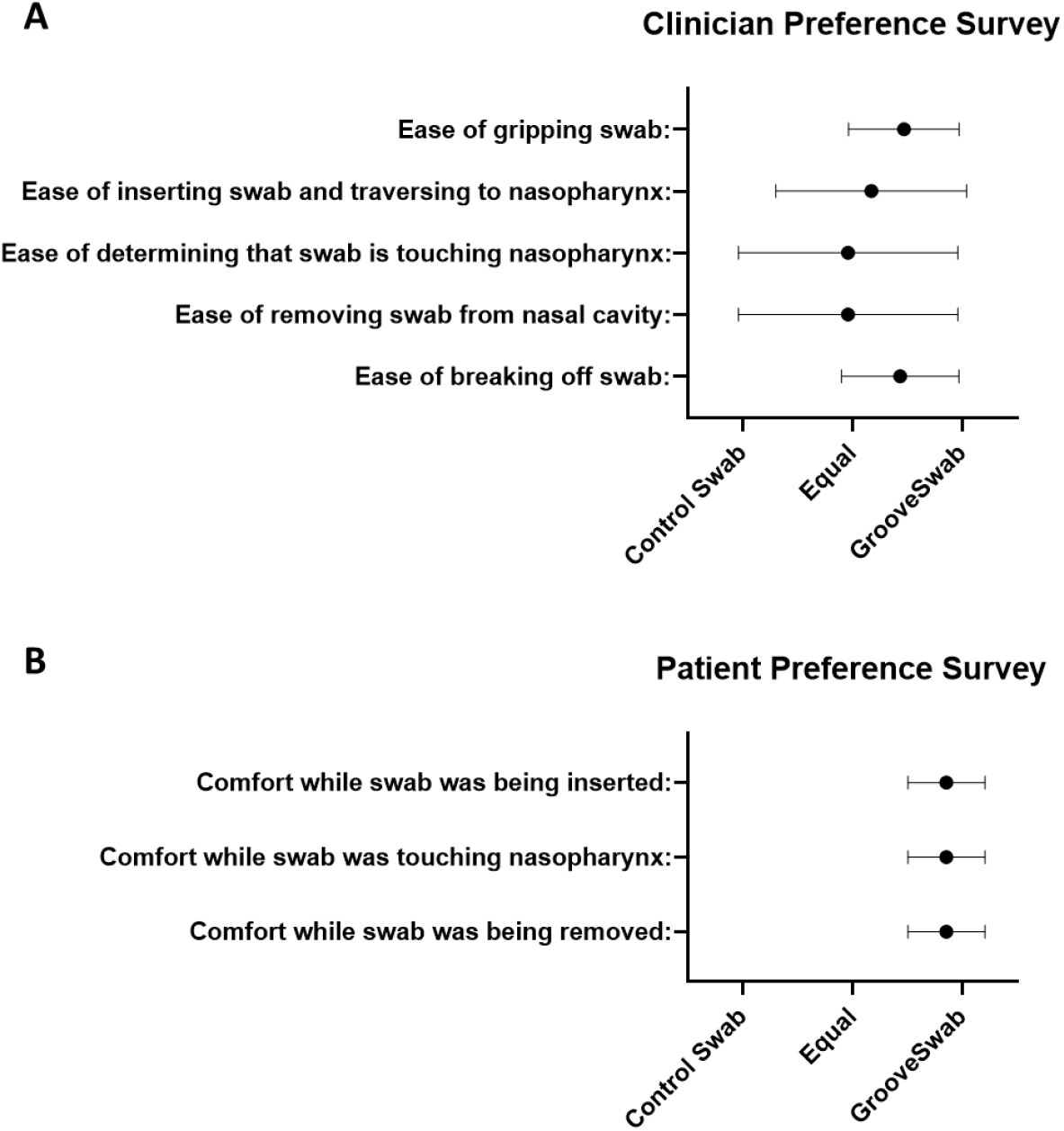
Survey results of clinician (A) and patient (B) preferences across various metrics for GrooveSwabs vs. control swabs showing preference for the GrooveSwab.

## DISCUSSION

Our results demonstrate the development of a nasopharyngeal swab composed of a single material (polypropylene) that can be manufactured in a single step using conventional, low-cost, injection molding technique that is available world-wide. Unlike traditional absorbent swabs, the GrooveSwab design uses a stacked ring structure inspired by microscale features of a cat’s tongue^14^ to enable both efficient collection and release of viscous mucus. This approach has the advantage of simpler manufacturing without requiring post-processing steps to add absorbent flocking coatings or components. Compared to 3D printed swabs, injection molded swabs can be manufactured at higher throughput and lower cost with robust quality control and regulatory processes readily available from decades of medical injection molding expertise in industry and regulatory bodies, which exist world-wide.

We found the unflocked GrooveSwab to perform comparably (>90% positive agreement threshold) to commercial swabs in a clinical trial. In a subset of inpatients recovering from COVID-19, the GrooveSwab detected SARS-CoV2 RNA in 3 cases that were not detected by a commercial control swab. In addition, clinician feedback in both trials indicated equivalence of the GrooveSwab to commercial control swabs, and usability surveys in a subset of patients also showed a significant preference for the GrooveSwab. We hypothesize that this may be due to the lack of absorption of mucus while transiting the nasal cavity, resulting in lower friction between polypropylene and the epithelium.

The rapid release capability of GrooveSwabs allows for more flexible clinical diagnostic scenarios, including temporary elution of samples into collection media by immersion into a tube and avoiding the need to later remove the swab head during analysis. Importantly, the non-absorbent head minimizes sample dilution by allowing for elution fluid volumes to be on the scale of tens to hundreds of microliters instead of milliliters while not impacting reproducibility. This may be valuable for assays without amplification steps, such antigen testing. Furthermore, this may support dry transport of collected specimens and avoid the need for collection media altogether. Additional validation would be required, but we have already demonstrated a high degree of stability and diagnostic performance in an anterior nares swab format of the GrooveSwab^15^.

The fluorescence assay using mock mucus samples we used offers a faster and more clinically-relevant test of sample collection and release properties of swabs compared to water absorption gravimetric measurements and absorption of low-viscosity water-based liquids spiked with known concentrations of nucleic acids PCR targets. The respiratory sample mimic substrate may be safer, more reproducible, and more accessible to researchers than clinically collected samples from healthy volunteers. However, a limitation of the porcine mucin-based mock mucus used here is a difference in microrheological properties that have been shown to differ in reconstituted mucus substrates compared to native respiratory tract mucus^16,17^. Standardization of mock mucus substrates may be helpful for comparing sample collection efficiency across NP and other swab products in development.

The simple, injection molded, GrooveSwab presented here demonstrates an effective means of achieving clinical diagnostic goals with a design optimized for both clinical and manufacturing needs. The non-absorbent collection head addresses supply chain challenges that accompanied the SARS-CoV2 pandemic and may offer advantages over absorbent swabs by improving patient comfort and enabling sample concentration. The simplicity of the biocompatible design may allow for scalable collection of diagnostic specimens from other sampling sites, including the airways, oral cavity, and wounds, which enables preparation for surveillance and monitoring of future disease outbreaks.

## Data Availability

Raw data not included in the manuscript are available from the corresponding author upon reasonable request.

## ACKNOWLEDGEMENTS

We are grateful to Drs. Jose Caro, Cody Callahan, and Ramy Arnaout for their helpful feedback on swab design features. This work was supported with funding from the Wyss Institute for Biologically Inspired Engineering at Harvard University.

## AUTHOR CONTRIBUTIONS

MRM, DEI, and RN conceived the swab design. DP, JT, and ICC provided manufacturing, regulatory, and clinical trial design support. RM, EG, WF, EF, SJS, DME, LG, HC, TKM, NM, CAUG, and JL designed and executed clinical studies. MRM, CS, DEI, and RN wrote the manuscript. All authors edited and reviewed the final manuscript.

## REFERENCES

1. COVID-19 Map. Johns Hopkins Coronavirus Resource Center https://coronavirus.jhu.edu/map.html.

2. Irving, S. A., Vandermause, M. F., Shay, D. K. & Belongia, E. A. Comparison of Nasal and Nasopharyngeal Swabs for Influenza Detection in Adults. Clin. Med. Res. 10, 215–218 (2012).

3. Iwasaki, S. et al. Comparison of SARS-CoV-2 detection in nasopharyngeal swab and saliva. J. Infect. 81, e145–e147 (2020).

4. Tu, Y.-P. et al. Patient-collected tongue, nasal, and mid-turbinate swabs for SARS-CoV-2 yield equivalent sensitivity to health care worker collected nasopharyngeal swabs. medRxiv 2020.04.01.20050005 (2020) doi:10.1101/2020.04.01.20050005.

5. Zou, L. et al. SARS-CoV-2 Viral Load in Upper Respiratory Specimens of Infected Patients. New England Journal of Medicine (2020) doi:10.1056/NEJMc2001737.

6. Smieja, M. et al. Development and Evaluation of a Flocked Nasal Midturbinate Swab for Self-Collection in Respiratory Virus Infection Diagnostic Testing. J. Clin. Microbiol. 48, 3340–3342 (2010).

7. Jamal, A. J. et al. Sensitivity of Nasopharyngeal Swabs and Saliva for the Detection of Severe Acute Respiratory Syndrome Coronavirus 2. Clin. Infect. Dis. doi:10.1093/cid/ciaa848.

8. Callahan, C. J. et al. Open Development and Clinical Validation of Multiple 3D-Printed Nasopharyngeal Collection Swabs: Rapid Resolution of a Critical COVID-19 Testing Bottleneck. J. Clin. Microbiol. 58, (2020).

9. Alghounaim, M. et al. Low-Cost Polyester-Tipped 3-Dimensionally-Printed Nasopharyngeal Swab for the Diagnosis of Severe Acute Respiratory Syndrome-Related Coronavirus 2 (SARS-CoV-2). J. Clin. Microbiol. (2020) doi:10.1128/JCM.01668-20.

10. Bennett, I. et al. The Rapid Deployment of a 3D Printed Latticed Nasopharyngeal Swab for COVID-19 Testing Made Using Digital Light Synthesis. medRxiv 2020.05.25.20112201 (2020) doi:10.1101/2020.05.25.20112201.

11. Ford, J. et al. A 3D-printed nasopharyngeal swab for COVID-19 diagnostic testing. 3D Print. Med. 6, 21 (2020).

12. Fahy, J. V. & Dickey, B. F. Airway Mucus Function and Dysfunction. N. Engl. J. Med. 363, 2233– 2247 (2010).

13. CDC. Information for Laboratories about Coronavirus (COVID-19). Centers for Disease Control and Prevention https://www.cdc.gov/coronavirus/2019-ncov/lab/guidelines-clinical-specimens.html (2020).

14. Noel, A. C. & Hu, D. L. Cats use hollow papillae to wick saliva into fur. Proc. Natl. Acad. Sci. 115, 12377–12382 (2018).

15. Pettit, M. E., Boswell, S. A., Qian, J., Novak, R. & Springer, M. Accessioning and automation compatible anterior nares swab design. J. Virol. Methods 294, 114153 (2021).

16. Lock, J. Y., Carlson, T. & Carrier, R. L. Mucus models to evaluate the diffusion of drugs and particles. Adv. Drug Deliv. Rev. 124, 34–49 (2018).

17. Huck, B. C. et al. Macro-and Microrheological Properties of Mucus Surrogates in Comparison to Native Intestinal and Pulmonary Mucus. Biomacromolecules 20, 3504–3512 (2019).

