## Supplementary figures and images for "Low cost, injection molded, nasopharyngeal swabs for addressing global diagnostic supply shortages"

### Supplemental Figure 1

**Material: FHR P5M4R**

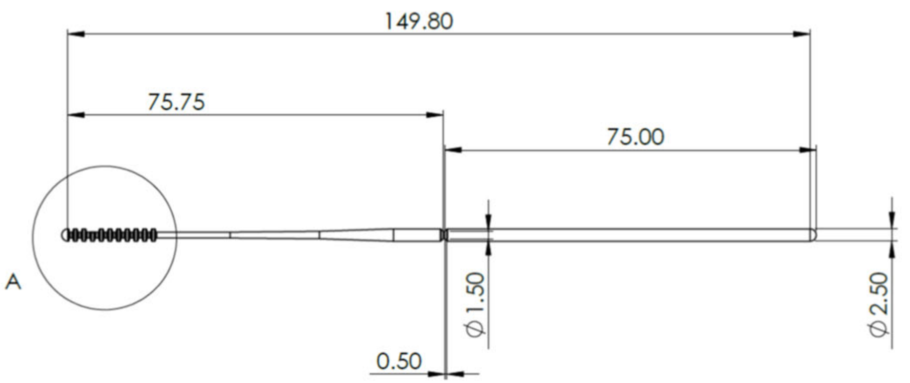


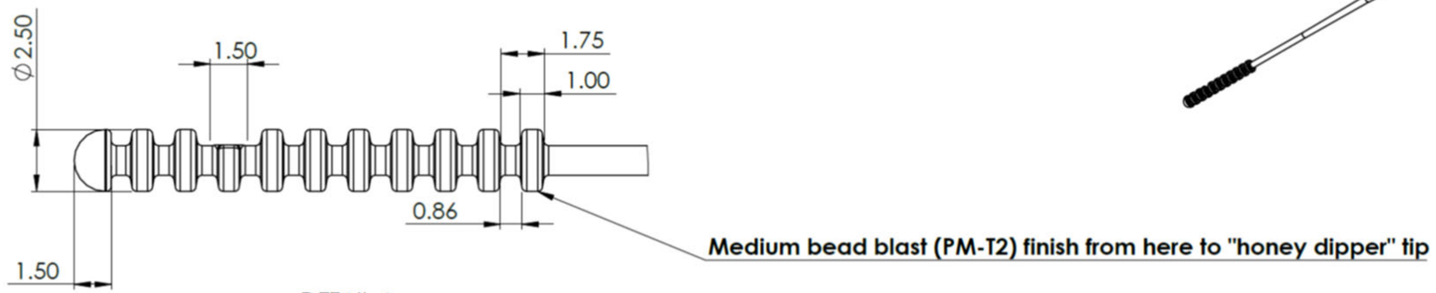


DETAIL A
SCALE 5 : 1

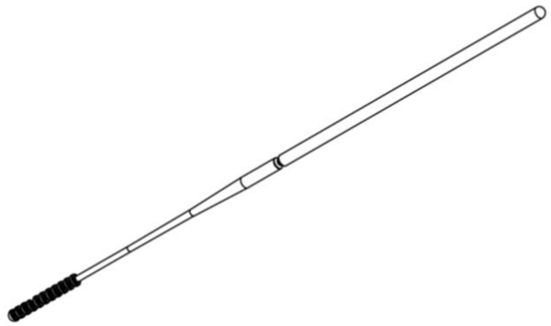
